# Short Telomeres and a T-Cell Shortfall in COVID-19: The Aging Effect

**DOI:** 10.1101/2021.05.19.21257474

**Authors:** James J. Anderson, Ezra Susser, Konstantin G. Arbeev, Anatoliy I. Yashin, Daniel Levy, Simon Verhulst, Abraham Aviv

**Affiliations:** School of Aquatic and Fishery Sciences, University of Washington, Seattle, Washington, 98195 USA; Department of Epidemiology, Mailman School of Public Health, Columbia University, New York, NY 10032, USA; New York State Psychiatric Institute, New York, NY, 10032 USA; Biodemography of Aging Research Unit, Social Science Research Institute, Duke University, Durham, North Carolina, 27705 USA; Population Sciences Branch, National Heart, Lung, and Blood Institute, National Institutes of Health, Bethesda, Maryland, 27705 USA; The Framingham Heart Study, Framingham, Massachusetts, 01702 USA; Groningen Institute for Evolutionary Life Sciences, University of Groningen, Groningen, The Netherlands; The Center of Human Development and Aging, New Jersey Medical School, Rutgers University, Newark, New Jersey, 07103 USA

**Keywords:** Aging, COVID-19, Lymphopenia, T cells, Telomeres

## Abstract

The slow pace of global vaccination and the rapid emergence of SARS-CoV-2 variants suggest recurrent waves of COVID-19 in coming years. Therefore, understanding why deaths from COVID-19 are highly concentrated among older adults is essential for global health. Severe COVID-19 T-cell lymphopenia is more common among older adults, and it entails poor prognosis. Much about the primary etiology of this form of lymphopenia remains unknown, but regardless of its causes, offsetting the decline in T-cell count during SARS-CoV-2 infection demands fast and massive T-cell clonal expansion, which is telomere length (TL)-dependent. We have built a model that captures the effect of age-dependent TL shortening in hematopoietic cells and its effect on T-cell clonal expansion capacity. The model shows that an individual with average hematopoietic cell TL (HCTL) at age twenty years maintains maximal T-cell clonal expansion capacity until the 6th decade of life when this capacity plummets by more than 90% over the next ten years. The collapse coincides with the steep increase in COVID-19 mortality with age. HCTL metrics may thus explain the vulnerability of older adults to COVID-19. That said, the wide inter-individual variation in HCTL across the general population means that some younger adults with inherently short HCTL might be at risk of severe COVID-19 lymphopenia and mortality from the disease.

**Significance Statement:** Declining immunity with advancing age is a general explanation for the increased mortality from COVID-19 among older adults. This mortality far exceeds that from viral illnesses such as the seasonal influenza, and it thus requires specific explanations. One of these might be diminished ability with age to offset the development of severe T-cell lymphopenia (a low T-cell count in the blood) that often complicates COVID-19. We constructed a model showing that age-dependent shortening of telomeres might constrain the ability of T-cells of some older COVID-19 patients to undertake the massive proliferation required to clear the virus that causes the infection. The model predicts that individuals with short telomeres, principally seniors, might be at a higher risk of death from COVID-19.

## Introduction

Transient lymphopenia is a common feature of acute viral respiratory infections (1). The drastic and prolonged lymphopenia of COVID-19, however, is distinctive and largely stems from falling counts of T cells (2-5). Regardless of the still poorly understood primary causes of this T-cell lymphopenia (2,3), the decline in T-cell count in COVID-19 demands fast and massive T-cell clonal expansion, which is telomere length (TL)-dependent (6,7). As hematopoietic cell TL (HCTL) shortens with age (8), T-cells from some older adults might lack the clonal expansion capacity required to offset the development of COVID-19 lymphopenia or recover from it (9). These individuals, we hypothesize, are at a higher risk of developing COVID-19 T-cell lymphopenia and severe disease.

In absence of an acute infection, the T cell turnover is slow because of the relatively long biological half-lives of naïve T cells and memory T cells in the circulation, i.e., ∼ 5 years and ∼ 5 months, respectively (10). In the face of SARS-CoV-2 infection, however, diminished TL-dependent T-cell proliferative capacity in older adults could result in a shortfall between T-cell depletion and repletion (11). Moreover, the clearance of SARS-CoV-2 requires clonal expansion and differentiation of naïve T cells into SARS-CoV-2 antigen-specific effector/memory (henceforth memory) T cells (2,3,5). Short naïve T-cell telomeres might thus limit adaptive immunity against the virus even without infection-mediated T-cell lymphopenia. We therefore modelled the relationships of TL-dependent T-cell clonal expansion capacity with age and virtually examined its relation to COVID-19 mortality in the general population.

## Results

Note: Abbreviations, symbols and units used in the mathematical development of the model are presented in Box 1.

### Box 1.

Abbreviations, meanings, units and values

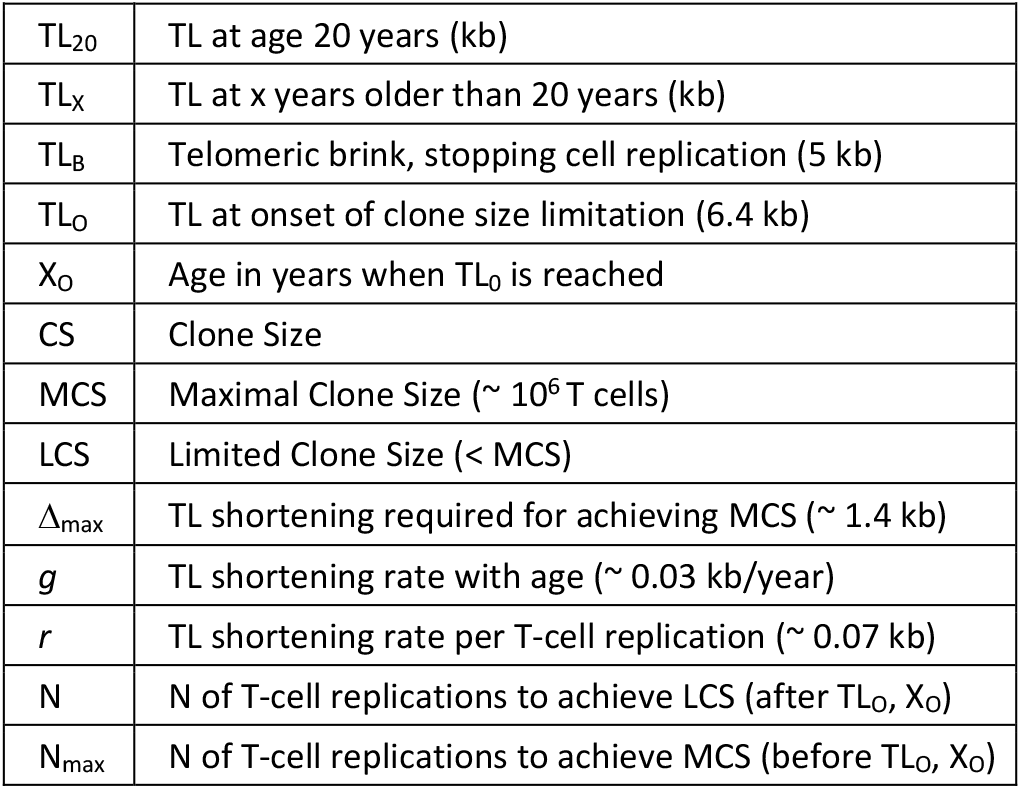

The following assumptions on T-cell replication and T-cell clone size (CS) drive our model (Fig. 1a): (i) T-cell TL-dependent cessation of replication is defined by a “telomeric brink” (TL_B_) that stops replication at 5 kilobases (8). (ii) TL of a naïve T cell at age 20 years (TL_20_) progressively shortens at a rate g of 0.03 kb/year (12,13) until it reaches the TL_B_. (iii) In exponential growth, i.e., 1→ 2→ 4→ 8→16, etc., a single naïve T cell can generate a maximal CS (MCS) of ∼ 10^6^ (one million) memory T cells through ∼ 20 replications (N_max_); this value was estimated based on the ∼ 1.4 kb TL difference between circulating naïve and memory T cells and the ∼ 0.07 kb telomere shortening per replication of cultured T cells (14). (iv) We denote the maximal TL shortening due to clonal expansion as Δ_max_ and the TL shortening per replication as r. (v) Due to age-dependent TL shortening, a naïve T-cell TL reaches the “telomeric onset” (TL_O_ = 6.4 kb) at “age of onset” (X_O_). Until X_O_, a naïve T cell can generate MCS. After X_O_, a naïve T cell can generate only a limited clonal size (LCS < MCS), as the TL of the clonal cells converges to the TL_B_. (vi) Most memory T cells are formed in response to new antigens during childhood and early adulthood (15), when HCTL is comparatively long (16-18), enabling the achievement of MCS.

**Fig. 1.**
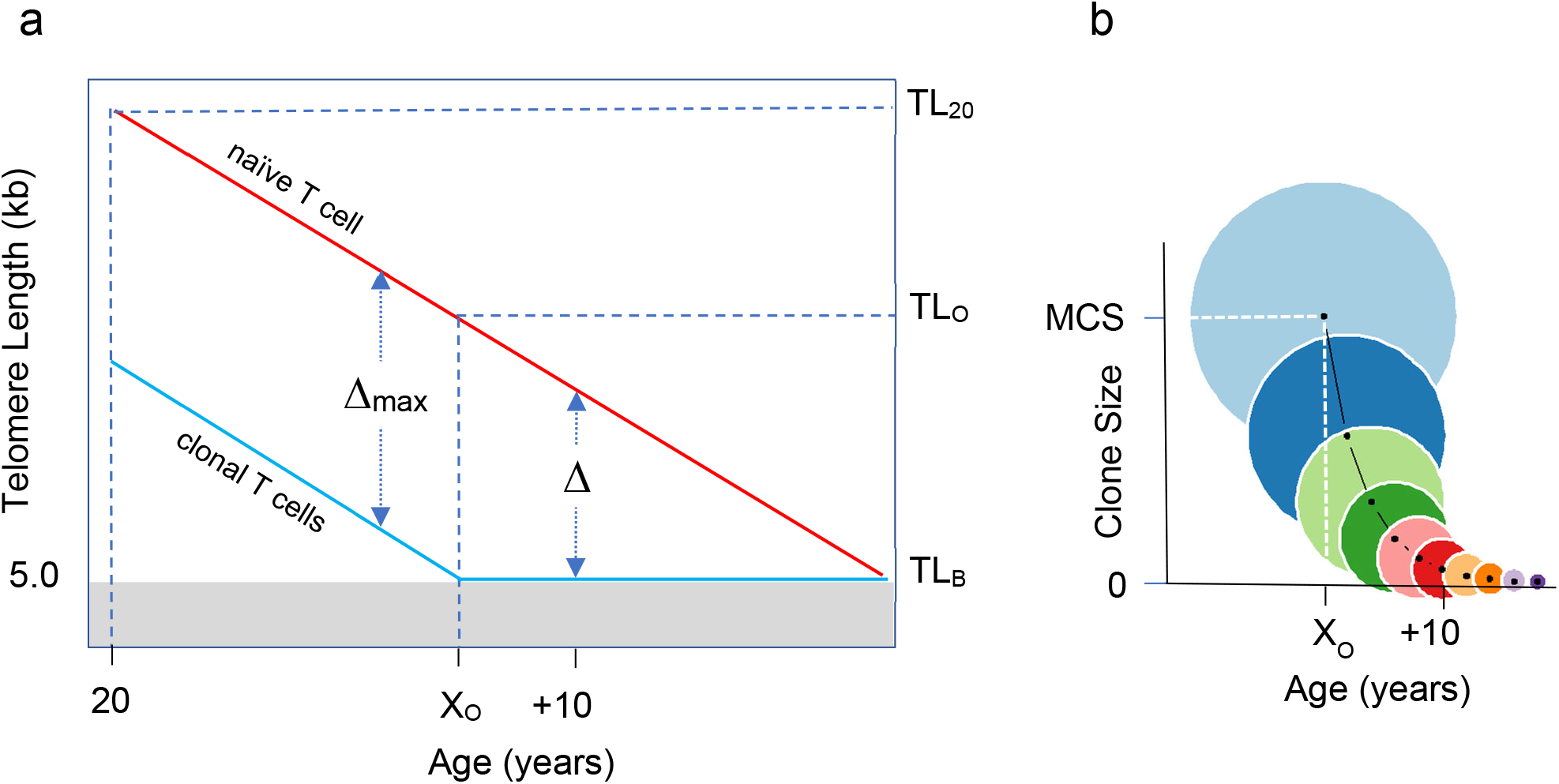
Age-dependent T-cell telomere length (TL) and its relation to T-cell clonal expansion. **a** displays age-dependent TL before 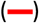 and after 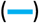 clonal expansion. Naïve T-cell clonal expansion shortens telomeres by Δ, where Δ_max_ is T-cell telomere shortening resulting from expansion to form the maximal clonal size (MCS). The telomeric brink (TL_B_) of 5 kb is TL that increases the risk of cessation of replication. TL_20_ is TL at 20 years, TL_O_ is telomeric onset, which indicates the shortest T-cell TL that enables attaining MCS. X_O_ is age of onset of clonal expansion limitation. **b** displays T-cell clonal expansion size vs age from onset X_O_. Circle areas depict relative clonal size in 2-year intervals after X_O_. Light blue circle is MCS.

We define CS by the number (N) of T-cell replications producing *CS* = 2^*N*^. As a clone expands, TL of its T cells progressively shortens, i.e., Δ= *rN*, where *r* is the telomere shortening due to T-cell replication. Prior to X_O_, the maximum number of T-cell replications in clonal expansion is *N*_max_ = Δ_max_/*r* = 20. After X_O_, the number of T-cell replications in clonal explansion is *N* = (*TL*_*X*_ − *TL*_*B*_)/*r*, where *X* designates age and the corresponding age-dependent TL of a naïve T cell is *TL*_*X*_ = *TL*_20_ − *g* (*X* − 20), where *g* is the TL shortening in the naïve T cell each year. The resulting X_O_ is the number of years it takes for a naïve T cell to reach TL_O_. These measures are defined:

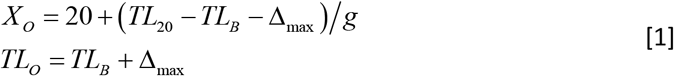

The clone size depends on age relative to X_O_ as follows:

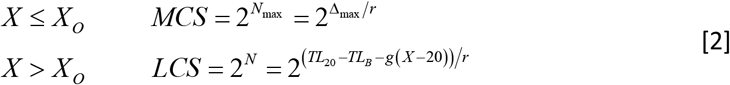

Note that after X_O_, while TL continues to shorten at its slow pace of 0.03 kb/year below TL_O_ of 6.4 kb (Fig. 1a), the clonal expansion capacity of T cells plummets exponentially. Consequently, in one decade after X_O_, the clonal expansion capacity of naive T cells is 5% of MCS (Fig. 1b).

The following considerations are relevant for appraising the model’s parameters and assumptions: First, while the MCS is based on the *in vivo* TL difference between naïve and memory T cells, the data on telomere shortening per T-cell replication are from cultured cells (14). A similar approach (based on data from circulating hematopoietic cells and telomere shortening in cultured cells) was previously used to generate consistent information on hematopoietic cell replicative kinetics (19,20). Second, the model’s TL parameters are derived from a large population-based study (8) that measured HCTL by Southern blotting (21). Its telomere data are consistent with another large-scale study that used Flow-FISH to measure HCTL (18). Third, the model is based on age-dependent shortening of HCTL and not T-cell TL. As TL differences among leukocyte lineages within the individual are far smaller than the inter-individual HCTL variation (22), T-cell TL largely mirrors HCTL. Fourth, the TL signal for cessation of cell replication originates from the shortest telomeres in the nucleus and not their mean TL (23,24). Using the Telomeres Shortest Length Assay (TeSLA), a method that tallies and measures the shortest telomeres (25), we recently showed that in patients with COVID-19 the shortest telomeres in peripheral blood mononuclear cells were associated with low lymphocyte counts (11). The principles that drive our model thus likely apply to the T cell’s shortest telomeres.

Consider now three individuals with average, long (one SD above the mean) and short (one SD below the mean) naïve T-cell TL_20_ (Fig. 2a) whose TL shortens with age (Fig. 2b): Up to X_O_ of 50 years, the naive T cells of the individual with the average TL_20_ can attain the MCS of ∼10^6^ cells. Thereafter, however, the ability of these naïve T cells to clonally expand declines in an exponential manner from the MCS to 0.43 x 10^6^, 0.24 x 10^6^ and 0.13 x 10^6^ cells at ages 53, 55 and 57 years, respectively (Fig. 2c). Next consider the individual with long T-cell TL_20_ (Fig. 2a). The ability of naive T-cells of this individual to achieve MCS extends to the X_O_ of 70 years (Fig. 2c). This suggests that even among older adults, some individuals can develop MCS when infected with SARS-CoV-2. In contrast, naïve T cells of the individual with short T-cell TL_20_ are able to achieve MCS only until the X_O_ of 30 years (Fig. 2c), inferring that some young adults might generate a poor T-cell response to SARS-CoV-2 infection. Our model thus suggests that in the overwhelming majority of the general population, naïve T cells can achieve MCS before early adulthood. Later, post X_O_, while age-dependent shortening of T-cell telomeres remains slow (Figs. 1a and 2b), it has a striking effect on clonal expansion of naïve T cells because the exponential nature of the process (Figs. 1b and 2c).

**Fig. 2.**
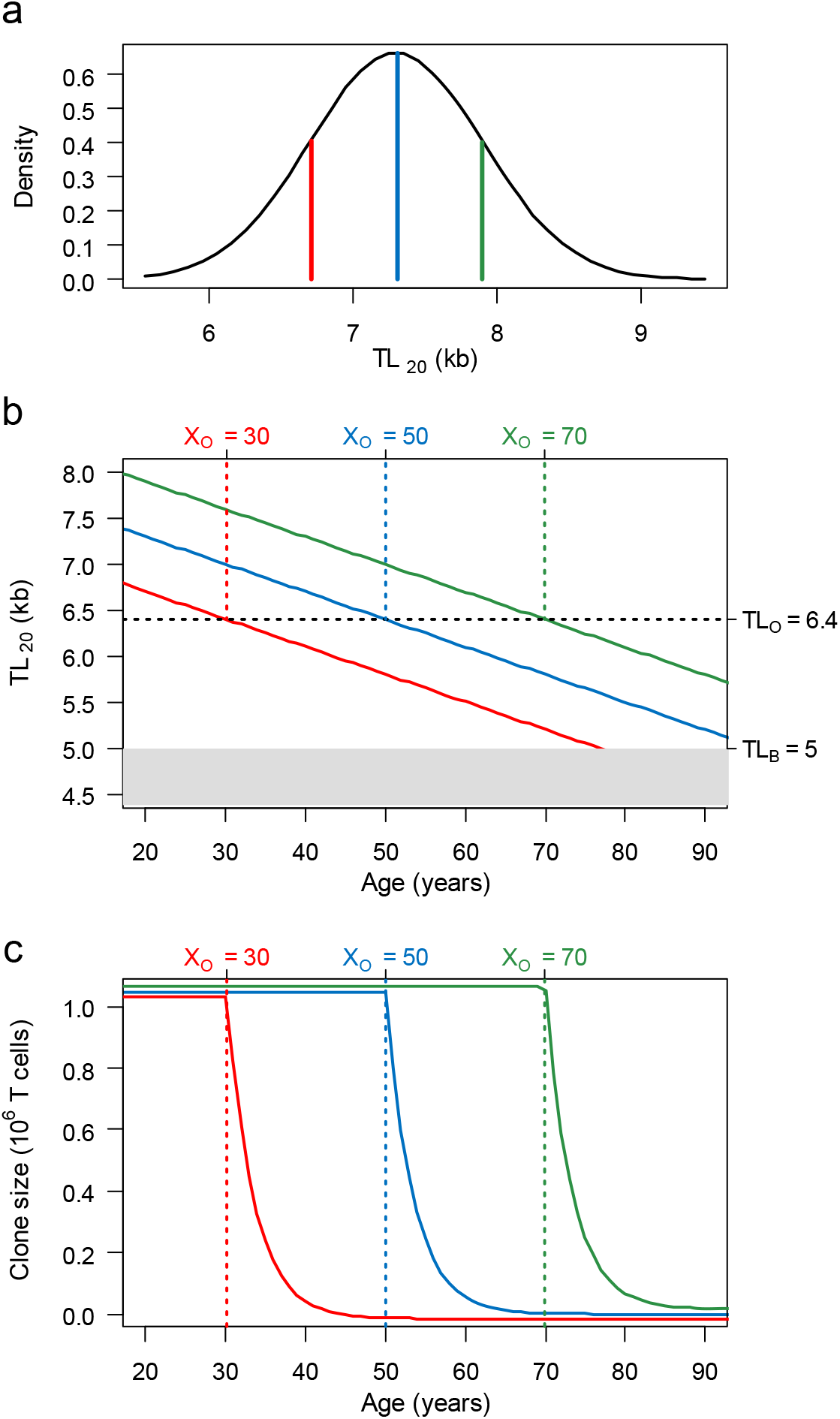
Population distribution of T-cell TL at age 20 (TL_20_), T-cell TL shortening with age, and age-dependent change in T-cell clone size (CS). **a** displays the TL_20_ distribution, showing mean TL = 7.3 kb 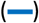, long TL (mean + SD) = 7.9 kb 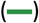, and short TL (mean – SD) = 6.7 kb 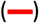. **b** displays age-dependent change in T-cell for mean, long and short TL_20_. Past the telomeric onset (TL_O_ = 6.4 kb), TL is insufficient to produce MCS because a full clonal expansion drops TL below the telomeric brink (TL_B_ = 5 kb). The TL_O_ is reached at different ages of onset (X_O_), i.e., an older age for T-cells with long T-cell telomeres and younger with T-cells with short telomeres. The age-dependent T-cell TL shortening (0.03 kb/year) for T cells with mean, long and short telomeres at TL_20_ is shown by the lines. **c** shows that the T-cell CS is partitioned by the X_O_ into plateau and slope regions. T cells with mean, long or short TL_20_ achieve MCS on the CS plateau, but their CS exponentially collapses (slope) once their TLs shorten below TL_O_ of 6.4 kb and exceed X_O_ (at different ages).

With aging, T cells of an increasing proportion of the population reach X_O_ (Fig. 3a). After reaching this TL landmark, LCS contracts rapidly to < 1% of MCS over 16 years. This rapid LCS contraction means that at any age, the individuals largely separate into two sub-populations: those with naïve T cells that can generate MCS and those with naïve T cells that can only generate LCS, which a few years after X_O_ is a small fraction of MCS. While naïve T cells of individuals in their twenties can overwhelmingly generate MCS, naïve T cells of most individuals in their seventies can at best generate clones that are typically less than one tenth of MCS (Fig. 3b).

**Fig. 3.**
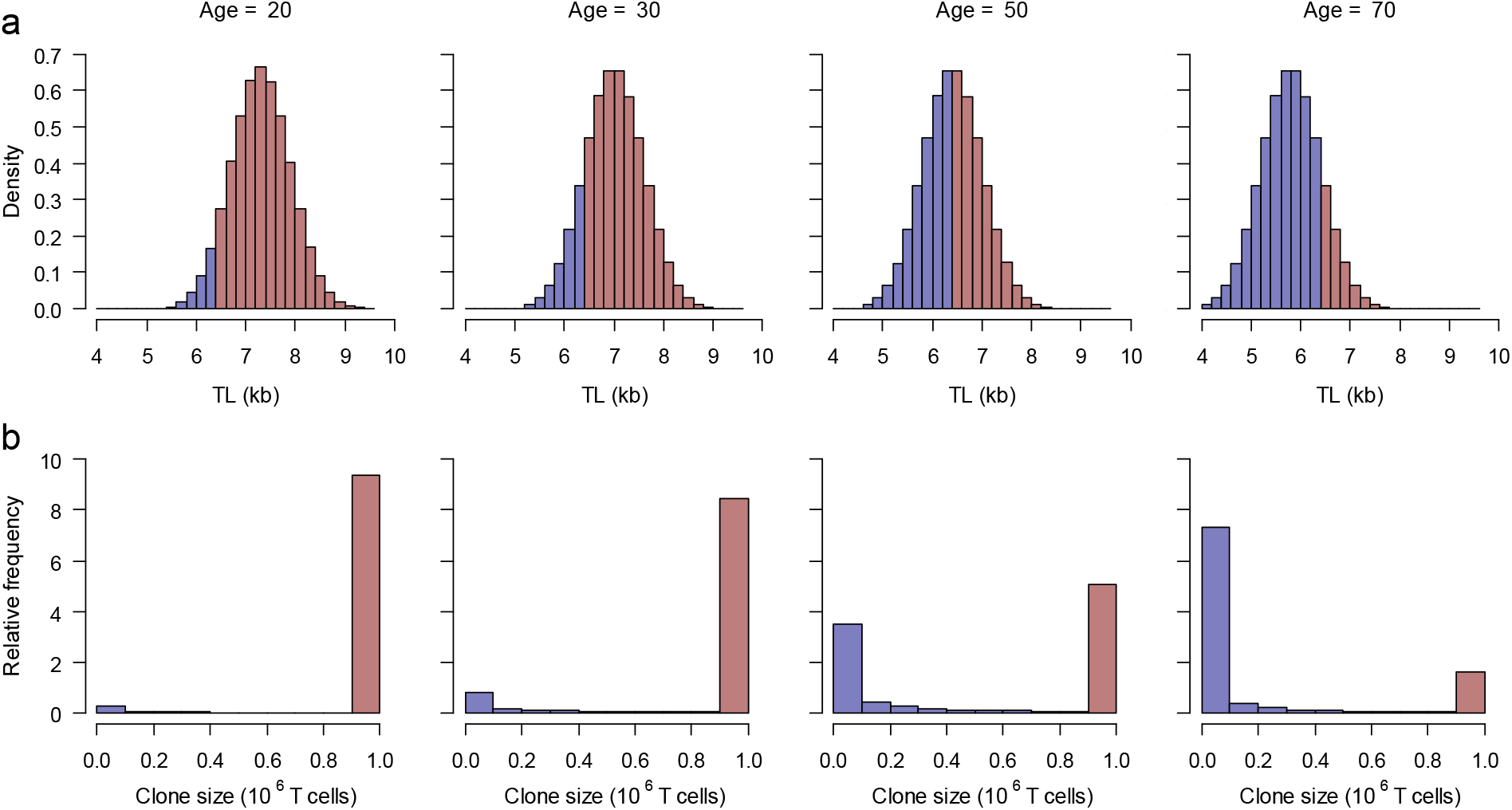
Shifts by age in naïve T-cell TL distribution and relative frequency (0 to 10) of T-cell clone size (CS) in the population. **a** displays the shift in TL_20_ distribution (Fig. 2a) resulting from age-dependent shortening of 0.03 kb/year. It depicts TL < TL_O_ (6.4 kb) by blue bars 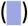 and TL > TL_O_ by red bars 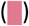. **b** displays relative frequency of CS’s generated by naïve T-cell clonal expansion corresponding to the categories of TL below or above TL_O_. It shows that maximal CS (MCS) of ∼ 10^6^ cells occurs in individuals with naïve T-cell TL > TL_O_, while limited CS (LCS) occurs in those with naïve T-cell TL ≤ TL_O_. At age 20, naïve T cells of nine out of ten individuals can generate MCS. At age 70, naïve T cells of less than two out of ten individuals can generate MCS, and seven out of ten generate clone sizes that are less than 0.1 MCS. At age 50 the population is approximately equally divided between the MCS and LCS groups.

What then might be the minimal TL-dependent T-cell CS that enables survival of an individual contracting COVID-19? The definitive answer awaits telomere and T-cell data in populations of COVID-19 patients. That said, we infer this CS based on Centers for Disease Control and Prevention (CDC) reports of the age-specific COVID-19 mortality rates for the United States (26) and the population size by age in 2019 (27). We computed an age-dependent hazards ratio of mortality relative to age 20 years (Hazards_20_) from COVID-19 and from general causes (27) other than COVID-19 (Methods). The Hazards_20_ yields an exponential increase of COVID-19 mortality with age, and for comparison, we also display Hazards_20_ for mortality from non-COVID-19 causes (Fig. 4a).

**Fig. 4.**
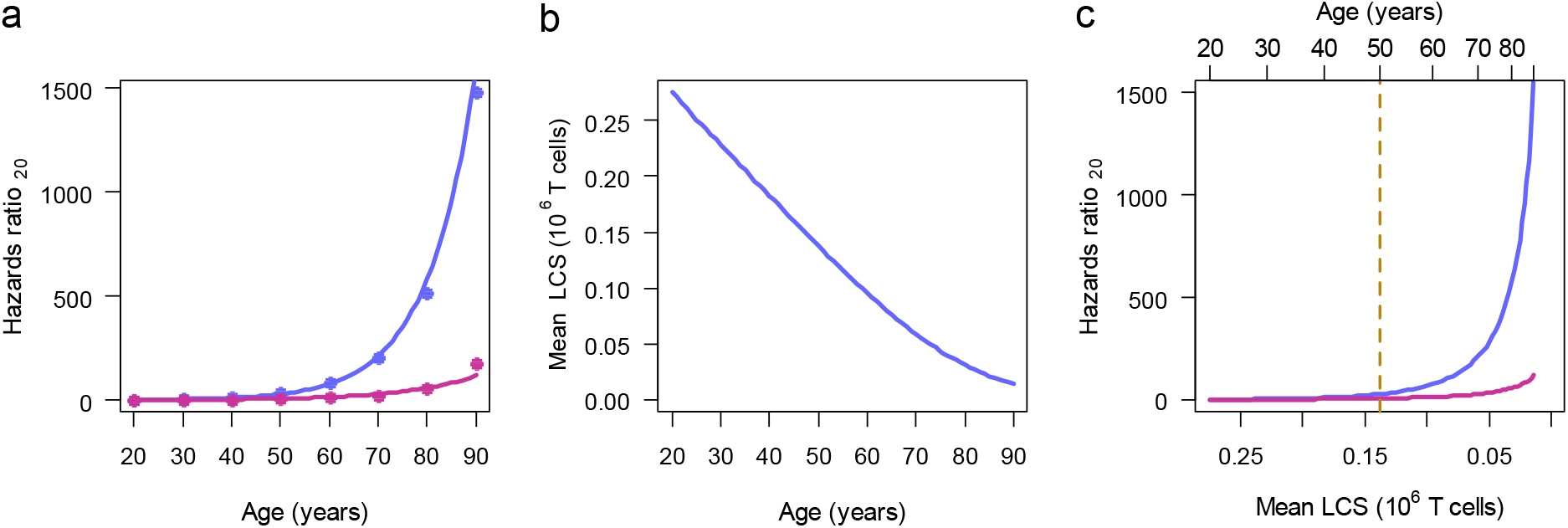
Steps linking mean limited clone size (LCS) to COVID-19 mortality and general mortality Hazards ratios_20_ in the population. **a** displays data based on COVID-19 mortality 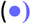 and non-COVID-19 mortality 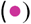, and corresponding exponential fitted relationships for Hazards ratios_20_ (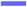 and 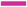). **b** displays the relationship of mean LCS in units of 10^6^ cells with age, generated with Eq. 2, using the TL_20_ distribution of Fig. 2a. **c** displays the relationships of Hazards ratios_20_ generated from COVID-19 mortality and non-COVID-19 mortality plotted against mean LCS obtained from **b**. The top of the panel also displays age. The divergence between the COVID-19 and non-COVID-19 mortalities occurs at mean LCS of ∼ 0.13 x 10^6^ T cells. At the corresponding age, 50 years, the population is about evenly divided into the LCS and MCS sub-populations (Fig. 3b). After this age, the majority of the population is in the LCS subpopulation, which is susceptible to COVID-19 mortality, whereas the MCS group is not.

We assumed that, as a subpopulation, individuals who generate MCS experience no T-cell TL-related COVID-19 mortality. We therefore calculated the mean CS for the LCS subpopulation only, i.e., adults older than X_O_ (Fig. 3a). The TL-limited clonal expansion of individuals in the LCS subpopulation, we assumed, might contribute to their propensity to die from COVID-19, given the association between lymphopenia and COVID-19 mortality. Fig. 4b shows that the mean LCS in adults older than X_O_ decreases in a near linear manner with age. Plots of the mean LCS vs. Hazards ratios_20_ of COVID-19 mortality and non-COVID-19 mortality suggests a divergence between the two trajectories during the 6^th^ decade (Fig. 4c). The figure also displays the mean LCS at the age of 50 years, which corresponds to the age at which the size of LCS subpopulation is similar to that of the MCS subpopulation (Fig. 3b). Mean LCS at this age amounts to ∼ 0.13 x 10^−6^ T cells.

The divergence of COVID-19 mortality from non-COVID-19 mortality when the mean LCS is about one tenth of MCS suggests the following: In the absence of COVID-19, ∼ 10% MCS is generally sufficient to accommodate the low turnover of T-cells (10). This LCS, however, might contribute to mortality in the face of SARS-CoV-2, because the infection demands massive T-cell clonal expansion to offset the primary cause of the dropping naïve T-cell count, and to generate memory T-cells that clear the virus.

Although our model is deterministic with fixed rates selected as best estimates from available literature, analyses reveals that uncertainty in the parameters and assumptions on the LCS cutoff definition have little effect on model conclusions. In *SI Appendix A1* an analysis of the propagation of error from *r, g*, TL_B_ and Δ_max_ indicate a 1.5% uncertainty in TL_O_ and ∼ 3.5 years uncertainty in X_O_ (Fig. S1a). The uncertainty in N and CS are significant prior to X_O_ but decline afterwards (Fig. S1b, c). However, the properties of the model are expressed in terms of the ratio of CS/MCS, so the model properties are independent of the absolute value of MCS.

The comparison of the COVID-19 and non-COVID-19 Hazards ratio_20_ (Fig. 4) assumes that COVID-19 mortality only occurred for CS < MCS. Correspondingly, other LCS cutoff levels, e.g., < 0.5 MCS or < 0.15 MCS, result in different relationships of LCS, hazards ratio_20_ and age. *SI Appendix A2* explores these relationships with alternative LCS cutoff levels. Alternative cutoffs have a minor effect on the percent of the population assigned to the LCS group (Fig. S2a) and therefore the cutoff has little effect on the distribution of the COVID-19-susceptible population with age as characterized by the bar colors in Fig. 3. However, the slope of mean LCS vs age declines with lower cutoff levels (Fig. 2b), which in turn compresses the hazards ratio_20_ curve vs the mean LCS (Fig. S2c). Consequently, the cutoff level changes the critical mean LCS at which the COVID-19 and non-COVID-19 hazards ratios_20_ diverge but this critical level is proportional to the cutoff level. Thus, a significant divergence of COVID-19 and non-COVID-19 hazards ratios_20_ is robust to the definition of the CS level susceptible to mortality.

## Discussion

With the exception of heritability (28), no other single factor so profoundly affects HCTL as does aging, explaining the key conclusion of our model. As SARS-CoV-2 memory T cells play a greater role than neutralizing antibodies in recovering from the infection (5), the aging effect on HCTL could impede adaptive immunity and heighten the risk for severe COVID-19. Moreover, we assume that MCS of ∼10^6^ cells applies not only for naïve T cells that clonally expand to produce memory T cells but also naïve T cells that clonally expand to produce naïve T cells. This means that regardless of the primary cause of COVID-19 T-cell lymphopenia, the T cell response after X_O_ will be compromised on two levels, i.e., formation of SARS-CoV-2-specific memory T-cells and replenishing the loss of naïve T-cells.

As illustrated in Figs. 2 and 3, our model might also apply to naïve T cells of a small subset of younger adults, whose HCTL is ranked at the lower range of the HCTL distribution in the general population (8,18). Comparatively short HCTL might also diminish naïve T-cell clonal expansion in response to SARS-CoV-2 infection in males, whose HCTL is shorter than in females from birth onwards (8,16,18,28), persons with atherosclerotic cardiovascular disease (29), obese persons and smokers (31,32), whose HCTL is respectively shorter than that in healthy, lean and non-smoking individuals. All these individuals with shorter HCTL are at a higher risk of severe COVID-19 and death from the disease (33-37).

Humans have comparatively short telomeres relative to their long lifespan (38,39), and therefore our model may not apply to most terrestrial mammals, including laboratory animal models that are used for viral research. For instance, TL-mediated replicative aging is probably not consequential during the 2-3-year lifespan of mice, considering their long telomeres (mean TL > 30 kb) and robust telomerase activity in their somatic cells. In contrast, the average human TL at birth is only ∼ 9.5 kb (16). As telomerase activity is repressed in replicating human somatic cells, their short telomeres experience further age-dependent shortening after birth. Although naïve T cells have some telomerase activity, it is insufficient to prevent their age-dependent telomere shortening, and aging may thus undermine the T-cell clonal expansion in many older humans. Whereas lymphopenia is a major prognostic feature of COVID-19 in older adults, when present, it is transient and of little prognostic value in children (40-42). The longer HCTL during early life potentially explain this clinical course.

Relatedly, the model shows that naïve T-cells with TL_20_ of one SD below the mean are unable to achieve MCS as early as after the third decade of life. This unexpected finding suggests that (a) under-estimation of naïve T-cell TL using population-based HCTL data, or (b) in response to pathogens, more naïve T cells might be tapped for clonal expansion (43) in young adults with short T-cell telomeres. Older adults may not have sufficient naïve T cells, particularly, naïve CD8 T cells, for this purpose (44).

We acknowledge the following limitations: The model draws on HCTL data from populations comprising principally whites of European ancestry in high-income countries. It should also be tested in populations of different ancestries and in low- and middle-income countries. The TL difference between naïve T cells and memory T cells likely reflects the clonal expansion in response to not only a single encounter but multiple encounters with a given antigen and its cross-reactive antigens. Thus, the MCS and LCS definitions in absolute T-cell numbers might be off the mark. Of note, however, the MCS and LCS can be expressed in the model in relative units of MCS (i.e., 0.5 MCS, 0.25 MCS, etc.) rather than absolute units (i.e., 0.5 x 10^6^, 0.25 x 10^6^ T cells, etc.), yielding identical results to those based on absolute T-cell numbers. Therefore, the principles of our model are likely to hold notwithstanding the above limitations.

In conclusion, the TL effect on naïve T-cell clonal expansion probably influences the outcome of SARS-CoV-2 infection. The insight generated by our model might set the stage for measurement of TL parameters not only in older adults but also the general adult population, helping to identify individuals vulnerable to severe COVID-19 because of short T-cell telomeres. These individuals might also be less likely to develop a lasting T-cell-mediated adaptive immunity in response to anti-SARS-CoV-2 vaccines. Finally, the ramifications of these conclusions go beyond the influence of TL on T-cell response to SARS Cov-2 infection and vaccination against the virus. They suggest that TL might be a limiting factor in immunotherapies whose efficacies depend on clonally expanding (in vivo and in vitro) transplanted hematopoietic cells, chimeric antigen receptor T cells, and tumor-infiltrating lymphocytes.

## Materials and Methods

The relationships between the age-dependent T-cell TL density and the relative proportion of CS in a population (Fig. 3) were derived from the distribution of HCTL at age twenty, extrapolated from age-specific density plots of HCTL (8). In Fig. 3a, the T-cell TL distribution for age 20 was derived from 1,000,000 random generations from a normal distribution with a mean = 7.3 kb and SD = 0.6 kb.

The combined male and female hazards ratios relative to age 20 in Fig. 4a were calculated from age-specific COVID-19-linked mortalities and total non-COVID-19 linked mortalities normalized by the age-specific US population. The hazards ratios, defined Hazards_20_ = (mortality_*age*_/population_*age*_)/(mortality_20_/population_20_), are based on the CDC records of 494,234 provisional COVID-19 deaths and 3,845,819 total deaths between January 1, 2020 through March 8, 2021 (26) and the 2019 US Census (27). Non-COVID-19 mortalities were estimated by subtracting the COVID-19 mortalities from the total mortalities for each age group.

R Code for generating the figures is provided in *SI Appendix B*.

## Data Availability

Data used in paper is available in supplemental material

## Acknowledgments

AIY and KGA research is supported by NIH grant RF1AG046860. SV human telomere research is funded NIH grant 5U24AG066528. AA telomere research is supported by NIH grants R01 HL134840, U01AG066529 and a grant from the Norwegian Research Council (NFR) ES562296.

## Disclaimer

The views expressed in this manuscript are those of the authors and do not necessarily represent the views of the National Heart, Lung, and Blood Institute; the National Institutes of Health; or the U.S. Department of Health and Human Services.

## Supplementary Information

### A1. Effects of parameter estimates on model response

Parameters (MCS, LCS, X_O_, TL_O_, g, r, TL_B_, TL_20_, and Δ_max,_ Box 1 in the paper) were derived from published data (Table S1 shows references). To demonstrate the contribution of these terms to the model uncertainty, we use the propagation of error approach assuming uncorrelated terms. Based on the equation *X*_*O*_ = 20 + (*TL*_20_ − *TL*_*B*_ − Δ_max_)/*g* the uncertainty in the TL_O_ measures is

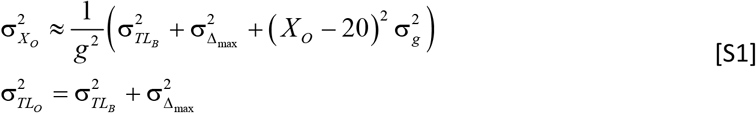

To estimate uncertainty of CS, first define the number of T-cell replications in clone expansion as *NX* = (*TL*20 − *g* (*X* − 20) − *TLB*)/*r* where age is X. Then variance in replications is

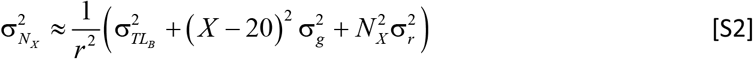

Next, to estimate uncertainty in CS, define clone size in terms of number of replications as 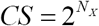, then the variance in the CS for a clone with limited expansion ability is

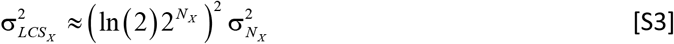

and the variance on MCS is calculated from equation (S3) by setting X to X_O_ giving

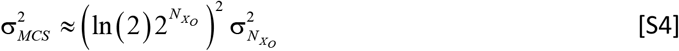

The uncertainties for these measures are calculated using the parameter estimates and standard deviations given in Table S1.

The uncertainty in the TL of the TL_O_ from Eq. [S1] is 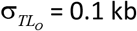 kb, or 1.5%. The uncertainty in X_O_ depends on TL_20_. Fig. S2a shows that the uncertainty varies slightly with TL_20_. Using the population mean TL_20_ of 7.3 kb the uncertainty is 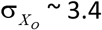 years.

The uncertainty in CS and number of replications depend on age X (Fig. S2 b, c). In the figures the uncertainty is normalized by measures prior to TL_O_, i.e., N_max_ = 20 and MCS = 2^20^. In panels b and c, the relative uncertainties decrease after the TL_O_, here set to X_O_ = 50 years to represent individuals with average TL_20_ of 7.3 kb. Notably, the large uncertainty in the MCS is wholly dominated by σ_r_ through Eq.[S2]. While the uncertainty in CS is significant, the properties of the model are expressed in terms of the ratio CS/MCS and not the actual value of MCS.

**Fig. S1.**
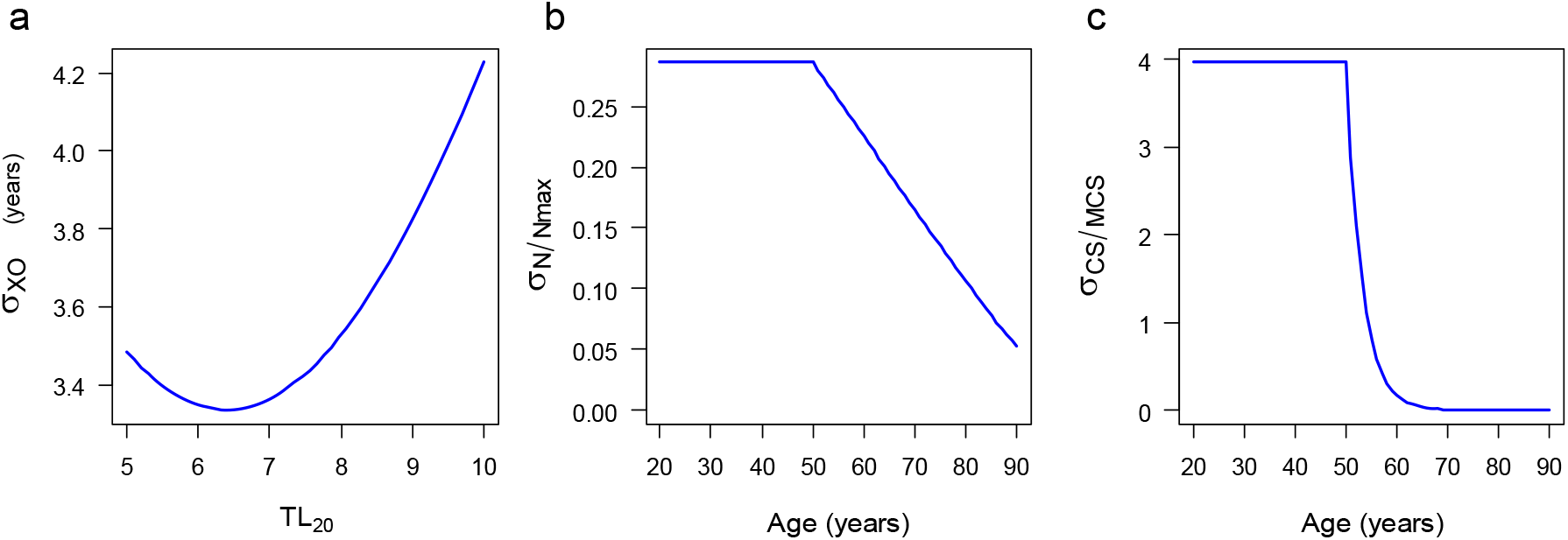
Uncertainty in parameter estimates. (**a**) Uncertainty in X_O_ with TL_20_. (**b**) Uncertainty in clone replication number (N) with age normalized by N_max_. (**c**) Uncertainty with age in CS normalized by the MCS.

**Table S1.** Parameter estimates and uncertainties.

| parameter | value | units | Reference |
| --- | --- | --- | --- |
| $\sigma_{\Delta_{max}}$ | 0.1 | kb | 1 |
| $\sigma_B$ | 0.0034 | kb | 2,3 |
| $\sigma_g$ | 0.00065 | kb/year | 1 |
| $\sigma_r$ | 0.02 | kb/replication | 3 |
| $\Delta_{max}$ | 1.4 | kb | 3 |
| $TL_B$ | 5 | kb | 1 |
| $TL_{20}$ | 5-9 | kb | 4 |
| $g$ | 0.03 | kb/year | 1 |
| $r$ | 0.07 | kb/replication | 1 |

### A2. Effect of limited clone size (LCS) definition on model response and correlation with observed mortality

The model assumes that mortality from COVID-19 only occurs when T-cell clonal expansion cannot achieve maximum clone size (MCS). We were unable to *a priori* select how LCS might affect mortality. We explored instead the relationship of mean LCS with years and the Hazards ratio_20_ for three cutoff levels used for defining LCS: (I) LCS < MCS, (II) LCS < 0.5 MCS, and (III) LCS < 0.15 MCS (Fig. S2). In cutoff I, any clone that cannot achieve a MCS is included in the LCS subpopulation and therefore susceptible to COVID-19 mortality. In cutoff II, only clones that are less than half the MCS are included in the LCS subpopulation and in cutoff III only clones that are less than 15% of the MCS are included.

The percent of the subpopulation in the LCS category (%LCS) increases from ∼ 5% at age 20 to ∼ 90% at age 90 (Fig. S2a). These percentages change little between the three cutoff levels. The maximum difference in % LCS in the definitions occurs at age 50 years. This close tracking with age indicates that the LCS cutoff has a minimal effect on the partition of the subpopulations in Figs. 3a and b. Note that the height of the histogram bars in Fig. 3b does not depend on the LCS cutoff.

Fig. S2b illustrates that the LCS cutoff has a strong effect on the mean LCS pattern with age. But for all cutoffs the mean LCS declines in a nearly linear manner with age. Thus, the assumption of a linear age-dependence of mean LCS with age is unaffected by the LCS cutoff choice.

Fig. S2c depicts the effect of the LCS cutoff on the Hazard ratio_20_. Here, because the cutoffs do not significantly affect linearity of mean LCS with age, they rescale the mean LCS at which the Hazards ratio_20_ of COVID-19 and non-COVID-19 diverge, which for Fig. 4c is taken as the critical Hazards ratio_20_ = 25.4. Then the mean LCS associated with this critical Hazards ratio_20_ linearly scales with the LCS cutoff as 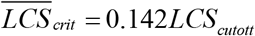.

**Fig. S2.**
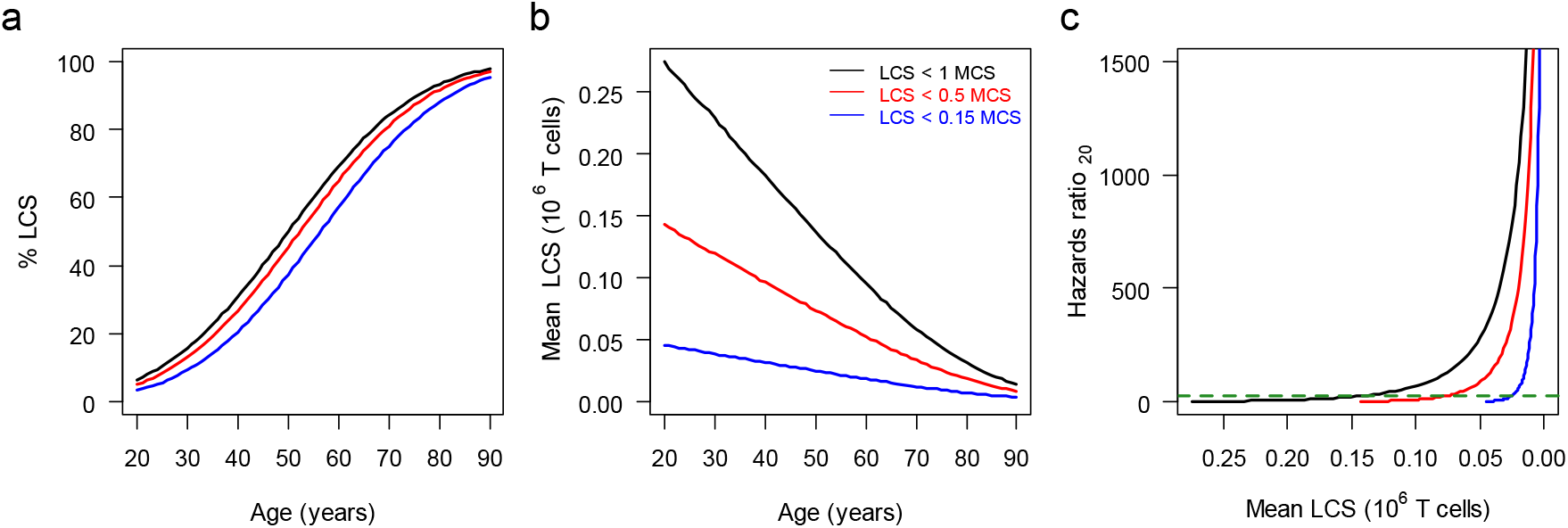
Model properties with three LCS cutoff levels: LCL < 1.0 MCS (**―**), LCL < 0.5 MCS 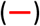 and LCL < 0.15 MCS 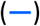. (**a**) % of population in LCS group by age for cutoff levels, (**b**) mean LCS vs age for cutoff levels, (**c**) Hazards ratio_20_ vs mean LCS for cutoff levels. Dashed line 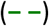 depicts Hazards ratio_20_ (25.4) at which COVID-19 and non-COVID-19 mortalities diverge.

